# Classifying dementia progression using microbial profiling of saliva

**DOI:** 10.1101/19004820

**Authors:** P. Bathini, S. Foucras, A. Perna, J.L. Berreux, M.A. Doucey, J.M. Annoni, L. Alberi Auber

**Affiliations:** Department of Medicine, Unit of Pathology, University of Fribourg, Fribourg, Switzerland; Neurology Clinic, Cantonal Hospital of Fribourg, Fribourg, Switzerland; Swiss Integrative Center for Human Health, Fribourg, Switzerland; Faculty of Medicine, University of Lausanne, Lausanne, Switzerland

**Keywords:** Alzheimer’s disease, olfaction, oral microbiome, cytokines

## Abstract

**Introduction:** There is increasing evidence linking periodontal infections to Alzheimer’s disease. Saliva sampling can reveal information about the host and pathogen interactions that can inform about physiological and pathological brain states.

**Methods:** A cross-sectional cohort of age-matched subjects (78) was segmented according to their chemosensory (University of Pennsylvania smell identification test; UPSIT) and cognitive scores (mini-mental score evaluation; MMSE and clinical dementia rating; CDR). Mid-morning saliva was sampled from each subject and processed for microbiome composition and cytokine analysis. Linear discriminant analysis (LDA) was used to unravel specific changes in microbial and immunological signatures and logistic regression analysis (LRA) was employed to identify taxa that varied in abundance among patients’ groups.

**Results:** Using olfaction we distinguish in the cognitively normal population a segment with high chemosensory scores (CNh, 27) and another segment with chemosensory scores (CNr, 16) as low as MCI (21) but higher than the AD group (17). We could identify stage-specific microbial signatures changes but no clear distinct cytokine profiles. Periodontal pathogen species as *F. villosus* decline with increasing severity of AD while opportunistic oral bacteria as *L. Wadei* shows a significant enrichment MCI. Conclusions: The salivary microbiome indicates stage-dependent changes in oral bacteria favoring opportunistic bacteria at the expenses of periodontal bacteria, while the inflammatory profiles remain mainly unchanged in the sampled population.

## 1. Background

Olfactory decline is an overarching preclinical sign of dementia and a strong predictor of impending neurodegenerative processes during aging [1]. Based on the low cost of the olfactory biometric, chemosensory testings are often implemented in the first medical screening along with memory testing. The University of Pennsylvania smell identification kit (UPSIT) called also “scratch and smell test” measures olfactory discrimination and identification, relying on central neural processing from the olfactory bulb to the olfactory cortices [2]. The olfactory circuitry is one of the first structures displaying a pronounced tauopathy already in healthy aging [3], and is one of the two central nervous system structures with direct access to the external environment representing an anatomical port of entry for neurotoxic species that can disseminate to the brain [1]. In support of the infectious hypothesis of Alzheimer’s disease (AD), a plethora of studies has accumulated evidence about a dysbiosis of microbial pathogens colonizing the head and neck area and their dissemination to the brain in the progression of AD [4]. In particular, *P. Gingivalis* a keystone periodontal pathogen and its antigens gingipains were found in the brain specimen from AD patients and its abundance correlated positively with the pathological progression of the disease [5]. Interestingly, levels of *P. Gingivalis* in saliva were inversely correlated to CSF in the AD subjects examined suggesting a potential evasion of the periodontal pathogens from the oral compartment to the brain. In this paper, we have used olfactory and cognitive scores to behaviorally segment the sampled population. Microbiome composition analysis indicates stage-specific changes but cytokine profiling indicates only subtle variations in pro-inflammatory chemokines. The present study supports the use of saliva as a monitoring biofluid for tracking the periodontal oral health in connection with brain aging.

## 2. Materials and Methods

The study was approved by the Swiss Ethics Board of the Canton of Vaud and Fribourg under the protocol n. CER-VD 2016-01627.

### 2.1. Study Participants

Participants were recruited at the Memory Clinic of the Cantonal Hospital of Fribourg. At the visit, subjects with cognitive impairment and their accompanying partner, serving as controls, provided written consent and all data collection was in compliance with the clinical protocol CER-VD 2016-01627 (selection criteria in Supplementary Table 1). All subjects were asked not to ingest food for at least two hours prior to the midmorning visit (9-11am). Before starting the test, they rinsed their mouth with an antiseptic solution, washed their mouth 4 times with water and then underwent cognitive testing (MMSE) and chemosensory probing (UPSIT, 16 odors [6]) taking typically 30 minutes. CDR assessment was done retrospectively based on the medical records or the interview with the certified clinical nurse (Table 1). Thereafter, about 2 ml of whole unstimulated saliva was sampled and processed as previously described [7].

**Table 1.** Classification of the cross-sectional study cohort.

| Variable | CNh | CNr | MCI | AD |
| --- | --- | --- | --- | --- |
| N (F) | 27 (16) | 15 (6) | 21 (13) | 17 (11) |
| Age | 67.0±9.2 | 68.1±10.0 | 73.2±8.1 | 71.1±6.6 |
| BMI | 26.3±5.0 | 25.2±4.6 | 24.6±4.6 | 26.0±4.1 |
| MMSE | 28.4±1.1 | 28.4±1.5 | 22.5±1.5<sup>‡</sup> | 14.2±4.3<sup>‡</sup> |
| CDR | 0.0±0.2 | 0.1±0.2 | 0.8±0.3<sup>‡</sup> | 1.4±0.6<sup>‡</sup> |
| UPSIT(%) | 0.7±0.1 | 0.5±0.1<sup>‡</sup> | 0.5±0.2<sup>‡</sup> | 0.3±0.2<sup>‡#</sup> |
| APOEε4 | 10 | 5 | 10 | 8 |
| Education * | 1.5±0.6 | 2.1±0.8 | 1.6±0.72 | 1.7±0.7 |

Values are means± standard deviations. F= female; M= male. MMSE= minimal mental score evaluation; UPSIT= University of Pennsylvania smell identification test. \* is for level of education: 1 = < 12 years of school; 2 = 12-15 years of school; 3 = > 15 years of school. <sup>‡</sup> p < 0.01 from pairwise comparison with CNh using Student's T-test. <sup>#</sup> p = 0.05 from pairwise comparison with MCI using Student's T-test.

### 2.2. APOE genotyping and 16S amplicon sequencing

Purified DNA from saliva (Norgen Biotech, Denmark) was genotyped for ApoE variants (ε2, ε3,ε4) using multiplex tetraprimers amplification according to a published protocol [8]. For the microbiome composition, V3-V4 16S rRNA amplicon sequencing was performed on an Illumina MiSeq platform, using standard protocols. The raw sequence data are available on request. The estimation of bacterial diversity within and between samples was performed with qiime2 version qiime2-2019.4 [9]. The Illumina BaseSpace 16S metagenomics tool was used to characterize the bacterial species, and their abundance, in the patients’ samples.

### 2.3. Bead-based immunoassay

15 *µ*l of saliva supernatant were probed using an inflammatory cytokines detection kit (Human Inflammation 16-Plex Panel, Aimplex, USA). Samples were run on the BD LSRII flow cytometer (BD Biosciences, San Jose, CA). The data were evaluated using the open access flow cytometry data analysis software (http://flowingsoftware.btk.fi/). Outliers were excluded in the final analysis.

## 3. Results

### 3.1. Clinical diagnosis

Participants are of European origin, representative of both genders, age-matched, with comparable BMI and same level of education. In the cohorts, the ApoEε4 allele distribution is aligned with the ApoEε4 incidence in Europeans (χ^2^=1.01, df=1, p=0.5)[10]. The subjects were segmented in groups according to their cognitive (MMSE and CDR) and olfactory scores (UPSIT; Table 1), of which MMSE and UPSIT are positively correlated (Supplementary Figure 1, A). In the cognitively normal group, we identify 2 populations, one with high olfactory scores (CNh=cognitively normal healthy) and one other with hyposmia (CNr=cognitively normal at risk; *t*=7.6, p<0.001) (Supplementary Figure 1, B). Mild cognitive impaired (MCI) subjects identified based on their MMSE/CDR scores (Table 1) also report hyposmia (*t*=6.7, p<0.001). Subjects with an MMSE below 20 and CDR larger than 1 are classified as AD and display the most severe olfactory impairment of all groups (*t*=8.9, p<0.001).

### 3.2. Oral microbiome analysis

Bacterial diversity does not differ among groups (Supplementary Figure 1, C) Nevertheless, linear discriminant analysis (LDA), applied to the oral bacterial content, reveals group-specific microbiome signatures (Figure 1, A) but less clear inflammatory profiles, based on the sampling of 16 cytokines of the innate and adaptive immunity (Figure 1, B and Supplementary Table 2). The cladograms (Supplementary Figure 2, A, C and E) represent the phylogenetic relationship between the bacterial taxa, which vary in abundance in the pairwise comparison between the CNh group and the CNr, MCI and AD groups. The linear discriminant analysis effect size (LEfSe) shows a depletion of bacterial taxa in MCI as compared to the other conditions (Supplementary Figure 2, B, D and F) and a systematic difference in Filifactor species between CNh and the other conditions. Indeed, Filifactor Villosus (*F. Villosus*) progressively decreases from CNr to AD (F_3,74_=3.5, p<0.05), whereas Leptorichia Wadei (*L. Wadei*) increases in abundance from CNr to MCI and then decays in AD (F_3,74_=5.1, p<0.01). In addition, Porphyromonas Gingivalis (*P. Gingivalis*) declines in MCI as compared to CNh (U=376, p<0.001) (Figure 1, D). Species being depleted in MCI as compared to CNh are Prevotella Tannerae (*P. Tannerae*; U=358, p=0.05), Filifactor Alocis (*F. Alocis*; U=399, p=0.05) and Cardiobacterium Valvarum (*C.Valvarum*; U=177, p=0.05) (Figure 1, E). Correlation analysis conducted for each group show the positive interactions among *P. Gingivalis* and Filifactor species (CNh, MCI and AD) and between *P. Gingivalis* and *L. Wadei* (CNr)(Supplementary Figure 3).

**Figure 1.**
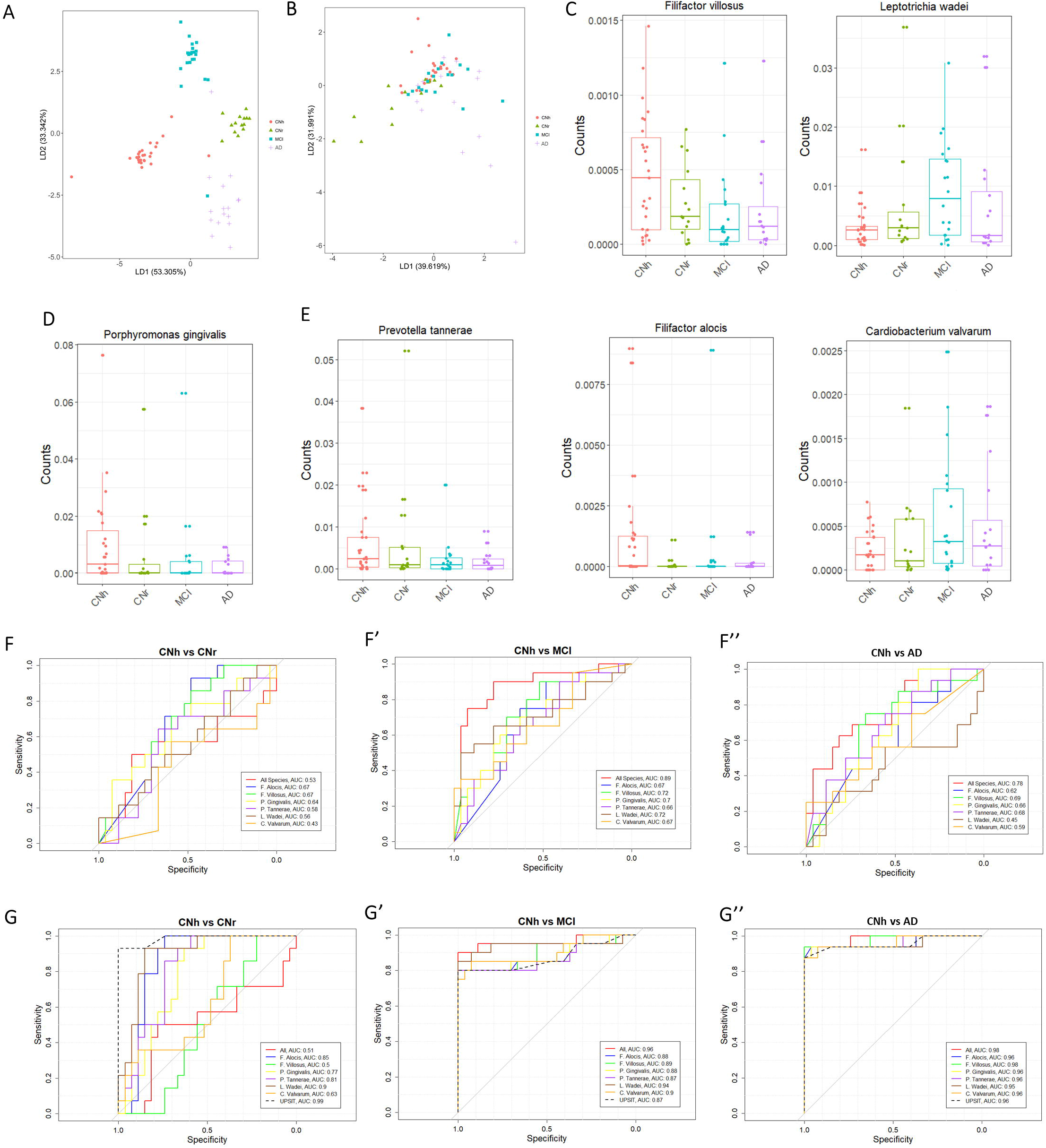
Microbiome and Inflammation profiling of saliva. **A)** LDA plots indicate that the oral microbiome composition is stage-specific. **B)** LDA analysis does not reveal any stage-specific inflammatory signatures. **C)** *L. Wadeii* and *F. Villosus* diverge significantly in CNr, MCI and AD as compared to CNh. **D)** *P. Gingivalis* is reduced in MCI as compared to CNh. **E)** *P. Tannerae*, *F. alocis* and *C. Valvarum* are significantly changed in AD as compared to CNh and tend to diminish in CNr and MCI. **F)** Logistic regression using species alone or together shows the discriminating power of DE species for CNh versus the other conditions. **G)** Logistic regression using a mixed model (DE species + UPSIT, and all DE species+ UPSIT=AII) improves the differentiating capacity. LDA=Linear Discriminant Analysis; DE= differentially expressed.

### 3.3. Predictive value of microbial signatures models

Logistic regression analysis test the predictive values of microbial species that differentiate alone or in aggregate the clinical groups (CNr, MCI, AD) as opposed to controls (CNh), considering MMSE as the dependent variable. In the distinction CNh versus CNr, *F. Alocis* and *F. Villosus* are the best differentiators of the conditions (Figure 1, F) with highest sensitivity (0.76 and 0.62) but modest specificity (0.44 and 0.55) and surpass the aggregate species model (Figure 1, F and Supplementary Figure 4). In the distinction CNh versus MCI, *F. Villosus* and *L. Wadei* are the best differentiators (Figure 1, F’) with moderate sensitivity (0.68 and 0.62) and specificity (0.55 and 0.59), while the aggregate model of all species show the best discriminating capacity with the highest sensitivity (0.72) and specificity (0.66). In the comparison, CNh versus AD, *F. Villosus* and *P. Tannerae* are the best differentiators (Figure 1, F”) with a moderate sensitivity (0.63 and 0.65) and moderate specificity (0.56 and 0.53). Also in this case, the aggregate model of all species show the best discriminating capacity with moderate sensitivity (0.67) and specificity (0.60). The mixed model of *L. Wadei* combined with UPSIT strongly differentiate CNr, MCI and AD as compared CNh (Figure 1, G-G”) with comparable sensitivity (0.76, 0.75 and 0.78) and moderately high specificity (0.63, 0.68, 0.66).

## 4. Discussion

Olfactory deficit is an overarching preclinical sign of dementia [11]. In our study olfactory phenotyping helped identifying subjects at risk, who retained high cognitive performance but were differentiated based on their chemosensory scores. These participants displayed an olfactory deficit comparable to MCI, while AD cases showed the strongest smell deficit, likely aggravated by their cognitive impairment [1]. Our operating framework, supports that CNr is likely an asymptomatic clinical condition preceding MCI and AD. Along with the behavioral phenotyping, in an effort to discover novel non-invasive diagnostic methods, we have employed salivary microbiome profiling with cytokine fingerprinting to assess pathogen-host interaction differentiating and staging AD. Previous work has shown that saliva can be employed as an accessible biofluid for biomarkers diagnostic of neurological conditions. A most recent study has indicated that in addition to Amyloid-β [12] and phosphorylated Tau species [13], which are detectable in the saliva severe patients, the antimicrobial humoral factor, lactoferrin, declines with the progression of AD [14]. This data strongly suggests a microbial dysbiosis with dementia progression. Our study using LDA confirms that despite taxa diversity remains unchanged, whereas there are specific changes in oral bacterial species, which are stage-dependent. LEfSe shows the strongest depletion in periodontal bacteria in MCI, while Leptorichia species tent to increase from CNr to MCI. Leptorichias are opportunistic bacteria found in oral biofilm and saliva. Increased amounts of *L. Wadei* are associated to rheumatoid arthritis by stimulating the release of antimicrobial chemokines and proinflammatory mediators, IL-6 and IL-8 [15]. Despite non-significant, our analysis indicates an increase in IL-8 (2 folds) in CNr and IL-6 (1.8 folds) and MIP-α (6 folds) in MCI. On the other hand, the progressive decline in anaerobic periodontal Filifactor species [16] is accompanied by a concomitant depletion of keystone periodontopathic pathogens *P. Gingivalis* and *P. Tannerae*. A decrease in *P. Gingivalis* counts was previously observed in the saliva from AD patients [5]. The depletion in periodontal taxa suggests either a reduction in number of teeth, due to earlier periodontal disease, or an oral dysbiosis with the advancement of opportunistic pathogens in AD subjects. The differentially expressed oral bacterial species alone or in association with UPSIT hold a moderate to strong predictive value in differentiating between clinical conditions (CNr, MCI and AD) as compared to healthy controls (AUC=0.67-0.96). Overall, this pilot work indicates that oral microbial signatures may be employed in larger studies as non-invasive classifiers of dementia alone or in addition to olfactory phenotyping.

## Data Availability

Raw Data available on request

## Acknowledgments

This research was supported by the Swiss National Foundation (SNF-163470; LA) and EKSAS Fellowship from the Swiss Ministry of Education (PB). We thank Muriel Jaquet and Eva Martin for technical assistance. We acknowledge the help of the undergraduate students who contributed to this project: Martina Minoli, Giulia Casal and Laura Leuenberger.

